# Wearable Prompt: In-Context Learning for Depression and Anxiety Prediction from Consumer Smart Ring Metrics

**DOI:** 10.64898/2026.07.31.26359400

**Authors:** Saeid Azadifar, Alireza Sameh, Maisa Niemelä, Vahid Farrahi

**Affiliations:** Research Unit of Health Sciences and Technology University of Oulu Oulu, Finland; Institute for Sports and Sport Sciences TU Dortmund University Dortmund, Germany

**Keywords:** Large Language Models (LLMs), Digital Phenotyping, Wearable sensing, Mental Health Prediction

## Abstract

Large language models provide a promising framework for wearable-based health prediction by converting structured physiological and behavioral measurements into natural-language prompts. In this paper, we investigate whether pre-trained lightweight open-weight LLMs can predict depression and anxiety symptoms from short-horizon consumer wearable data. Using 4–8 days of Oura Ring data from 1,285 participants in the Northern Finland Birth Cohort 1986, we convert activity, sleep, heart rate, heart rate variability, demographic, and anthropometric measurements into structured prompts. We evaluate Llama 3.1, BioMistral, and Qwen 2.5 under zero-shot, rule-based, and few-shot in-context learning settings. To contextualize LLM performance, we compare them against machine learning models and recurrent neural networks. Our results show that prompt design is critical for LLM-based wearable inference. Zero-shot LLMs achieve high accuracy but largely predict the majority class, failing to identify participants with depression and anxiety symptoms. In contrast, few-shot prompting substantially improves positive-class detection. Llama 3.1 with four in-context examples achieves the strongest performance, with 0.92 accuracy, 0.82 macro-F1, and 0.69 F1 for the positive class, among evaluated models. These findings suggest that lightweight LLMs can use in-context examples to better interpret structured wearable summaries and possibly provide a scalable direction for mental health prediction from consumer wearable data in combination with pre-trained LLMs.

**Code base:** https://github.com/saeidazadifar1988/OuraLLM

## I. Introduction

Consumer wearables provide a scalable and non-invasive opportunity to capture daily behavioral and physiological digital metrics, such as sleep, physical activity, inactivity, heart rate (HR), and heart rate variability (HRV) [1]. Recent research has demonstrated that data generated from wearables can be used to assess mental health [2]. Combined with machine learning (ML), these data enable digital phenotyping of mental health problems including depression and anxiety symptoms in free-living [3]. However, wearable data could be challenging to analyze because they are multivariate, noisy, partially missing, temporally dependent, and strongly influenced by individual differences. Wearable devices and metrics are of interest because they can potentially complement traditional assessments with passive, high-resolution data collected in free-living settings [1]. Yet, reliable predictive frameworks based on wearable time-series trajectories remain difficult to develop. Classical ML models often depend on hand-crafted features and feature engineering, while recurrent neural networks (RNN) such as LSTM and GRU can model temporal patterns but are still sensitive to short observation windows, imbalanced data and cohort heterogeneity. More recently, large language models (LLMs) have been explored for wearable health prediction by converting time-series summaries into natural-language prompts, but their performance remains highly dependent on prompt design and contextual grounding [4].

In this paper, we develop a predictive framework for depression and anxiety using Oura ring metrics from 1,285 participants from the Northern Finland Birth Cohort 1986 (NFBC 1986) 33-year follow-up. Depression and anxiety categories are defined using the Hopkins Symptom Checklist (HSCL-25). Our motivation is to examine how wearable-derived metrics and large language models (LLMs) can be jointly used for detecting depression and anxiety symptoms. This study is guided by two key questions: (1) To what extent can short-horizon wearable data collected over 4–8 days capture signals predictive of depression and anxiety and (2) whether in-context learning with LLMs can effectively use structured wearable metrics and achieve competitive predictive performance compared with traditional ML and sequence-based models. Fig. 1 illustrates the overview of the study protocol. This study makes the following contributions:

**Fig. 1:**
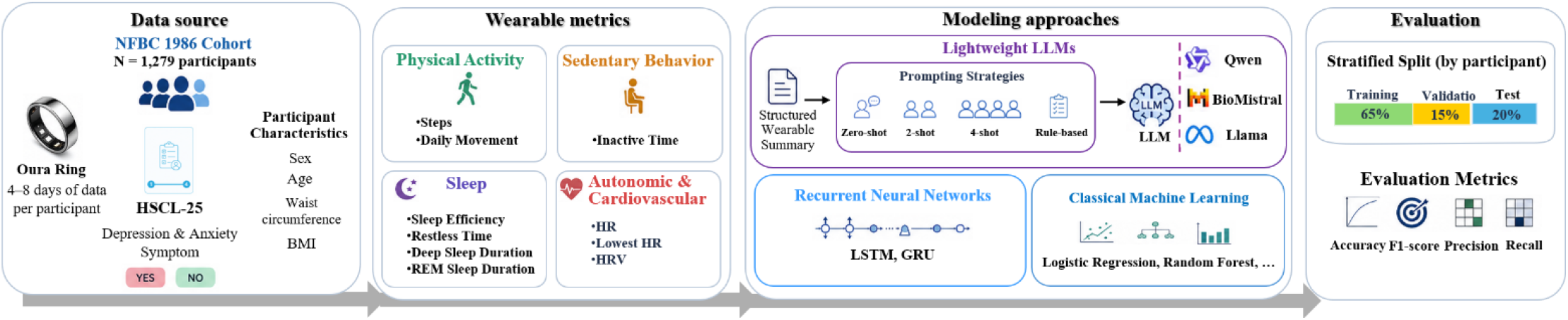
The overview of the study

- **Wearable-based mental health predictive framework:** We develop and evaluate predictive models for depression and anxiety using short-horizon (4–8 days) wearable data from a large, population-based cohort (NFBC 1986), capturing real-world physiological and behavioral signals.
- **Lightweight LLMs for physiological inference:** We investigate the effectiveness of lightweight, resource-efficient open-weight LLMs, leveraging in-context learning and other prompting strategies to use structured wearable summaries for predicting symptoms of anxiety and depression.
- **Systematic comparison of modeling paradigms:** We compare ML models, recurrent neural networks (LSTM and GRU), and large language models (LLMs) under the same experimental framework.
- **Insights for scalable multimodal digital health systems:** We provide empirical evidence on the strengths and limitations of LLM-based approaches for non-linguistic health data, highlighting their potential for scalable, real-world deployment in continuous mental health monitoring and intervention

## II. Related work

- Wearable- and smartphone-based mental health monitoring is closely connected with digital phenotyping, which uses. collected behavioral and physiological signals to characterize symptom-relevant changes in daily life [1]. Studies have shown that mobility, phone-use behavior, sleep-related patterns, and physical activity can provide meaningful insights into mental health and related outcomes [1]. Later research applied ML to engineered features derived from passive sensing, including activity regularity, sleep disruption, circadian regularity, and autonomic measures such as HR and HRV, to estimate or predict mental health states [5]. However, these feature-based approaches remain sensitive to missing data, class imbalance, small samples, limited behavioral variability, and cross-participant heterogeneity [1], [5]. In parallel, clinical time-series research has advanced sequence-aware modeling through RNN-based longitudinal risk prediction [6], representation learning for time-series prediction [7]. More recent work has extended this direction toward robust time-series learning, wearable- and ambient-sensor anomaly detection [8], and health prediction using LLM-based components [9]. These studies motivate models that can learn from temporal trajectories rather than relying only on hand-crafted summaries, while also underscoring the continued need for sufficient labeled longitudinal data and robust generalization across individuals.

LLMs introduce a different framework for supporting mental health risk assessment from wearable-derived data [10]. Rather than training a task-specific model on raw time-series data, structured metrics or time-series summaries can be converted into natural-language descriptions and provided to the model as prompts. This approach builds on in-context learning, where LLMs condition their outputs on task instructions or a small number of labeled examples at inference time, without updating model parameters [11]. Prompting strategies, including zero-shot and few-shot learning, have shown strong influence on performance in clinical NLP tasks [12].

In wearable health research, Models such as Health-LLM [4] convert sensor data into textual prompts for prediction tasks and other studies, such as ECG-Doctor explore LLM-based physiological interpretation [13]. However, most existing work has focused on general health prediction, activity recognition, ECG interpretation, or longer-horizon sensing data, and has often relied on relatively small or population-specific datasets, such as PMData with 16 participants and LifeSnaps with 71 participants [14]. Consequently, much less is known about whether lightweight open-weight LLMs can reliably predict depression and anxiety symptoms from short-horizon consumer wearable data, particularly when labeled mental health samples are limited.

## III. Methodology

### A. Dataset

Data for this study comes from the NFBC 1986 33-year follow-up [14]. Participants were asked to wear the second-generation Oura Ring (ōura Health, Finland) on any finger of their non-dominant hand. Depression and anxiety labels are defined using the HSCL-25, which is a validated tool for assessment of psychological distress including questions on both depression and anxiety symptom severity. Following previous studies, participants with HSCL-25 scores >1.75 were assigned to the positive class and others to the negative class. The final dataset included 1,279 participants, of whom 175 (13.7%) were positive and 1,104 (86.3%) were negative, indicating class imbalance.

Participants were included in this study if they had between 4 and 8 consecutive days of valid measurements. The dataset contains daily physiological and behavioral features, including step count, inactive time, daily movement, deep and REM sleep duration, sleep efficiency, HR, lowest HR, and HRV. We additionally incorporate demographic and anthropometric variables, including age, sex, body mass index (BMI), and waist circumference which were collected during clinical study visit.

### B. Problem Formulation

In this study, depression and anxiety prediction is formulated as a binary classification task. Let *D* = 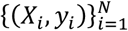 denote a dataset of *N* participants, where *X*_*i*_ is the multivariate wearable-derived time-series for participant *i*, and *y*_*i*_∈ {0,1} denotes the corresponding label indicating the absence or presence of depression and anxiety symptoms. Each participant is associated with a sequence of daily measurements collected over a time window where *T*_i_ ∈ [4,8]and *x*^(*t*)^∈ ℝ^*d*^ represents the feature vector at day *t*. The objective is to learn a predictive function *f* that maps wearable-derived signals to a probability estimate.

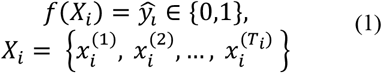

### C. Prompting Strategy, In-context Learning and LLMs

We converted each participant’s data into a structured natural-language input. This input included demographic and anthropometric variables and observation length, along with wearable time-series features, including steps, inactivity, movement, deep sleep, REM sleep, sleep efficiency, HR, lowest HR, and HRV. We evaluated four prompting strategies to assess the effect of in-context learning: zero-shot prompting, rule-based expert-driven prompting, few-shot prompting with two and four examples. The zero-shot prompt included only the task definition and the participant’s data, asking the model to directly predict whether the participant showed symptoms of depression or anxiety based on the wearable profile. The rule-based prompt added expert-informed heuristics related to activity, inactivity, sleep, HR, and HRV. The few-shot prompts further included labeled exemplar cases before the target participant, enabling inference-time adaptation without updating the model parameters. The prompt structures are summarized in Fig. 2.

**Fig. 2:**
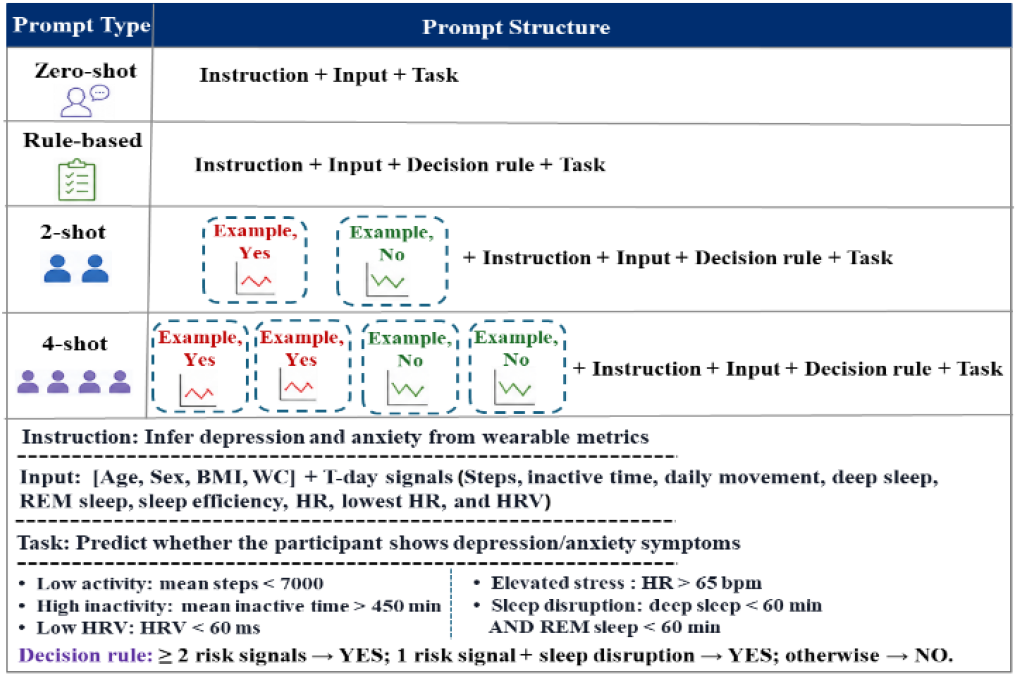
Overview of the prompting strategies

We evaluate three pretrained open-weight large language models as prompt-based classifiers: Llama-3.1-8B [15], BioMistral-7B [16], and Qwen2.5-7B [17] representing general-purpose, biomedical, and instruction-tuned LLMs.

## IV. Results

### A. Experimental Settings

All experiments were implemented in Python using standard ML and DL libraries, including scikit-learn, PyTorch, and the Hugging Face Transformers framework for LLMs. The experiments were conducted on a workstation equipped with 32 GB memory. The dataset was split into training (65%), validation (15%), and test (20%) sets, with a shared test set used for consistent evaluation. Few-shot exemplars were selected only from the training set to prevent information leakage. Classical ML models, including Logistic Regression (LR), Support Vector Machine (SVM), Random Forest (RF), and XGBoost, were trained on aggregated feature representations. Hyperparameters were tuned using grid search on the validation set. Recurrent neural network models, including LSTM and GRU, were implemented in PyTorch and trained using sequence inputs with appropriate padding for variable-length data.

### B. Main results

The models were evaluated using accuracy, precision, recall, and F1-score, with particular emphasis on recall and F1-score due to the imbalanced nature of the classification task. As shown in TABLE I substantially improved performance of LLMs for depression prediction compared with zero-shot and rule-based prompting strategies. Among all evaluated methods, Llama 3.1 with 4-shot prompting achieved the best overall performance, reaching 0.92 accuracy and 0.82 macro F1, while also providing the highest positive-class F1-score (0.69). In contrast, zero-shot prompting across all LLMs yielded high accuracy (0.86) but completely failed to identify positive cases (positive-class F1 = 0.0). BioMistral and Qwen 2.5 also benefited from few-shot prompting, achieving macro F1-scores of 0.66 and 0.63 with 4-shot prompting, respectively.

**TABLE 1:** Performance comparison of LLMs, RNNs, and classical ML models for wearable-based depression and anxiety prediction under different prompting strategies (Acc: Accuracy; Prec: Precision; Rec: Recall for the positive (YES) class; ZS: Zero-shot; RB: Rule-based; 2S: 2-shot prompting; 4S: 4-shot prompting)

| Model | Prompt | Acc | Prec | Rec | F1 | Macro-F1 |
| --- | --- | --- | --- | --- | --- | --- |
| LLM |  |  |  |  |  |  |
| Llama 3.1 | ZS | 0.86 | 0.00 | 0.00 | 0.00 | 0.46 |
|  | RB | 0.58 | 0.12 | 0.33 | 0.18 | 0.45 |
|  | 2S | 0.90 | 0.67 | 0.47 | 0.56 | 0.75 |
|  | 4S | **0.92** | **0.75** | **0.63** | **0.69** | **0.82** |
| BioMistral | ZS | 0.86 | 0.00 | 0.00 | 0.00 | 0.46 |
|  | RB | 0.58 | 0.12 | 0.33 | 0.18 | 0.45 |
|  | 2S | 0.67 | 0.13 | 0.25 | 0.17 | 0.48 |
|  | 4S | 0.88 | 0.62 | 0.29 | 0.39 | 0.66 |
| Qwen 2.5 | ZS | 0.86 | 0.00 | 0.00 | 0.00 | 0.46 |
|  | RB | 0.48 | 0.14 | 0.57 | 0.23 | 0.42 |
|  | 2S | 0.76 | 0.21 | 0.28 | 0.24 | 0.55 |
|  | 4S | 0.77 | 0.31 | 0.53 | 0.39 | 0.63 |
| RNN |  |  |  |  |  |  |
| LSTM | — | 0.49 | 0.16 | 0.66 | 0.26 | 0.43 |
| GRU | — | 0.72 | 0.12 | 0.17 | 0.14 | 0.49 |
| ML |  |  |  |  |  |  |
| LR | — | 0.86 | 0.33 | 0.06 | 0.10 | 0.51 |
| SVM | — | 0.86 | 0.00 | 0.00 | 0.00 | 0.46 |
| RF | — | 0.86 | 0.50 | 0.31 | 0.39 | 0.65 |
| XGBoost | — | 0.86 | 0.50 | 0.31 | 0.39 | 0.65 |

### C. Few-shot Improvement vs rule-based Prompting

Fig. 3 illustrates the effect of few-shot in-context learning. Performance gains were most pronounced for the minority “YES” class and macro F1, indicating improved detection of participants with depression and anxiety symptoms rather than reliance on majority-class prediction. Llama 3.1 showed the most consistent improvement, with 4-shot prompting outperforming 2-shot prompting across recall and F1 for the positive class. BioMistral and Qwen 2.5 also improved with additional examples, although the gains were less stable.

**Fig. 3:**
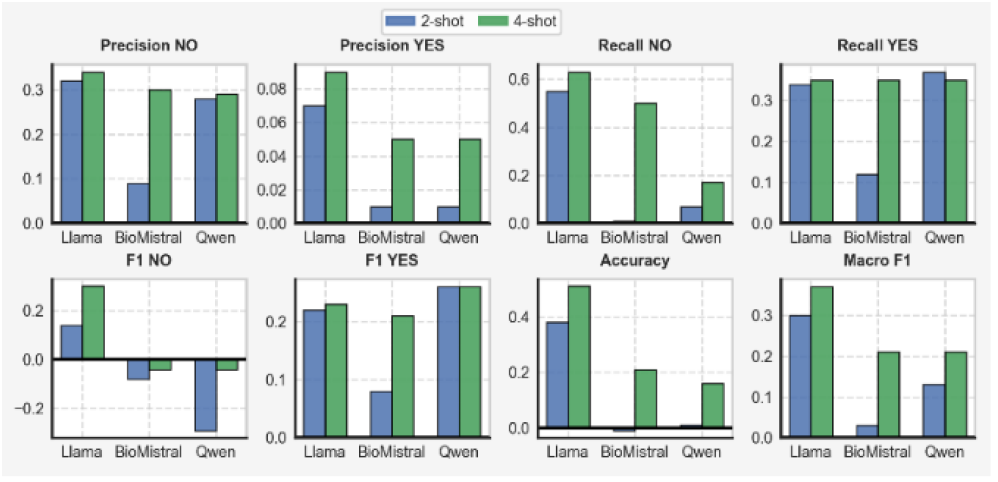
Performance improvement of 2-shot and 4-shot prompting over rule-based prompting across LLM frameworks, evaluated using precision, recall, F1-score, accuracy, and macro F1 metrics.

## D. Discussion

The results show that prompt-based LLMs can be effective for short-horizon wearable mental-health prediction, but their performance depends strongly on prompt context. Zero-shot prompting achieved high accuracy in some cases, but this was mainly due to majority-class prediction and weak detection of the depression and anxiety class. In contrast, few-shot prompting consistently improved minority-class performance, with Llama 3.1 using four examples achieving the best overall results. This suggests that in-context examples help LLMs better align wearable metrics with symptom labels. The weaker performance of rule-based prompting suggests that fixed thresholds alone cannot capture the multi-factorial nature of depression and anxiety signals. Overall, the findings suggest that LLMs are most useful when wearable summaries are paired with representative in-context examples, rather than used as zero-shot classifiers.

## V. Conclusion

In this study, lightweight open-weight LLMs were investigated for depression and anxiety prediction using short-horizon consumer wearable time-series data. We converted Oura Ring-derived physiological and behavioral metrics, participant’s demographic data, and anthropometric measurements into structured prompts and evaluated zero-shot, rule-based, and few-shot in-context learning strategies. Compared with classical ML and RNN baselines, few-shot LLMs showed stronger performance, particularly for detecting the minority positive class. Llama 3.1 with four in-context examples achieved the best results, indicating that labeled demonstrations help LLMs better interpret wearable-derived patterns and reduce majority-class bias. These findings suggest that prompt-based LLMs could serve as a flexible and scalable framework for wearable-based depression and anxiety symptom prediction, while also highlighting the need for careful prompt design, exemplar selection, and evaluation with class-sensitive metrics. Future work should examine longer sensing windows, retrieval-based exemplar selection, and multimodal models that combine time-series encoders with LLM reasoning.

## Data Availability

NFBC data is available from the University of Oulu, Infrastructure for Population Studies. Permission to use the data can be applied for research purposes via electronic material request portal. In the use of data, we follow the EU 395 general data protection regulation (679/2016) and Finnish Data Protection Act. The use of personal data is based on cohort participant's written informed consent at his/her latest follow up study, which may cause limitations to its use. Please, contact NFBC project center NFBCprojectcenter(at)oulu.fi) and visit the cohort website for more information.

## Acknowledgment

The research leading to this publication was co-funded by the European Union’s Horizon Europe Research and Innovation Programme under the Marie Skłodowska-Curie Actions grant agreement No. 101126602 (Data4Healthcare), and the University of Oulu. The present study is connected to the DigiHealth and 6GESS strategic profiling projects at the University of Oulu supported by the Research Council of Finland (project number 326291, 336449) and the University of Oulu. This study has also received funding from the Ministry of Education and Culture in Finland [grant numbers OKM/20/626/2022, OKM/76/626/2022, OKM/68/626/2023]. VF is supported by TU Dortmund university. The funders played no role in designing the study; collecting, analyzing, and interpreting the data; or writing the manuscript. NFBC1986 33-35-years follow-up study received financial support from University of Oulu (Strategic funding from donations) and Oulu University Hospital (K65760).

